# Within-family heritability estimates for behavioural and disease phenotypes from 500,000 sibling pairs of diverse ancestries

**DOI:** 10.1101/2025.09.17.25336022

**Authors:** Loic Yengo, Yanyu Liang, 23andMe Research Team, Xin Wang, Julie M. Granka, David M. Evans, Julia Sidorenko, Peter M. Visscher

## Abstract

Quantification of the direct effect of genetic variation on human behavioural traits is important for understanding between-individual variation in socio-economic and health outcomes but estimates of their heritability can be biased by between-family indirect genetic effects. In contrast, using within-family variation in DNA sharing is robust to most confounding factors including shared environmental effects and population stratification. Yet, accurate estimates for most traits are not available using this design, and none for non-European ancestry populations. Here, we analyse approximately 500,000 sibling pairs with diverse ancestries and obtain robust and precise heritability estimates for 14 phenotypes, including two well-studied model traits (height and BMI), five behavioural phenotypes and two common diseases. We find substantial heritability for smoking initiation (0.34 ± 0.05), alcohol consumption (0.18 ± 0.04), number of children (0.27 ± 0.11) and personality (“talk versus listen”, 0.48 ± 0.13). In addition, we estimated large heritability for two common diseases, type 2 diabetes (T2D: 0.43 ± 0.06) and asthma (0.34 ± 0.06), whose risk factors include behavioural traits. Overall, we show concordant estimates across ancestry groups and highlight a significant contribution of shared environmental effects for behaviour and T2D risk, which may have inflated between-family estimates. Altogether, our results demonstrate that substantial genetic variation underlies complex traits, common disease and exposures, that estimates are concordant across ancestries and that they are larger than has been accounted for by GWAS to date.

## INTRODUCTION

Heritability is a fundamental parameter that informs the extent to which trait differences between individuals are determined by genetic factors. Estimating heritability is critical for understanding the causes of the resemblance between relatives in disease risk and other traits, for predicting the effect of natural selection on phenotypes, and for quantifying the upper limit of polygenic risk prediction.^1^ Different designs have been proposed to estimate heritability, each having specific strengths and limitations.^2^ For behavioural traits, experimental designs based upon the correlation between relatives (e.g., twins) or genome-wide association studies (GWAS) are particularly prone to biases because of systematic between-family effects such as gene-environment correlation and assortative mating.^3–5^

Currently, the Full Sibling Identity-by-descent (IBD) Regression (FSIR) method is the gold standard for estimating heritability.^6^ This method uses variation in DNA sharing between full siblings, that is due to Mendelian segregation, to produce estimates that are robust to most confounders including shared environmental effects and population stratification. However, FSIR typically has lower statistical power compared to other designs and thus requires hundreds of thousands of sibling pairs to yield precise estimates. In 2018, an extension of the FSIR method was proposed by Young and colleagues^7^ to account for more complex family structures and was applied to estimate heritability of 14 quantitative traits in the Icelandic population. Their method requires extensive pedigree data, which is typically not available in biobanks and direct-to-consumer databases, and therefore their reported estimates from complex pedigrees have not yet been replicated elsewhere. Young et al.^7^ also showed results from FSIR, which we will use for comparison herein.

To date, the most powerful FSIR analyses used approximately 119,000 sibling pairs of European ancestry to generate robust heritability estimates for height and body mass index (BMI).^8^ Therefore, a substantial gap remains for most phenotypes (especially for behavioural traits and diseases) and for populations with other ancestries.

In this study, we use genomic data from up to 547,126 full sibling pairs of diverse ancestries to estimate the heritability (hereafter denoted h^2^, and its estimate 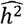) of 9 quantitative traits and 5 binary phenotypes, including 5 behavioural traits (smoking initiation, alcohol consumption, number of children, handedness and a personality trait) and 2 common diseases with behaviour-associated risk factors (asthma and type 2 diabetes).

## RESULTS

### Phenotypes and identity-by-descent (IBD) statistics

Descriptive statistics for all phenotypes are provided in **Supplementary Table 1**. We analysed binary phenotypes on an observed 0-1 scale and developed a new method to transform estimates to an underlying liability scale as described in **Supplementary Note 1** and **Supplementary Figures 1–4**. Untransformed heritability estimates for binary traits are also provided in **Supplementary Table 2**. Overall, we show highly concordant IBD sharing distributions between ancestry groups (**Supplementary Table 3**). Throughout the manuscript, statistical significance of 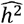 is called when the associated two-sided Wald-test p-value is lower than 0.05/42 1.2 × 10^−3^, which corresponds to the Bonferroni multiple-testing threshold for the 42 trait-ancestry pairs analysed. Estimates reported and discussed below were obtained using the recombination rate stratified FSIR method,^5^ but we also report estimates from unstratified analyses in **Supplementary Table 4**. Phenotypic correlations (after adjusting for covariates) between siblings for all traits are also reported in **Supplementary Table 4** and **Fig. 1**, which show remarkably concordant estimates across ancestry and ethnicity groups.

**Figure 1.**
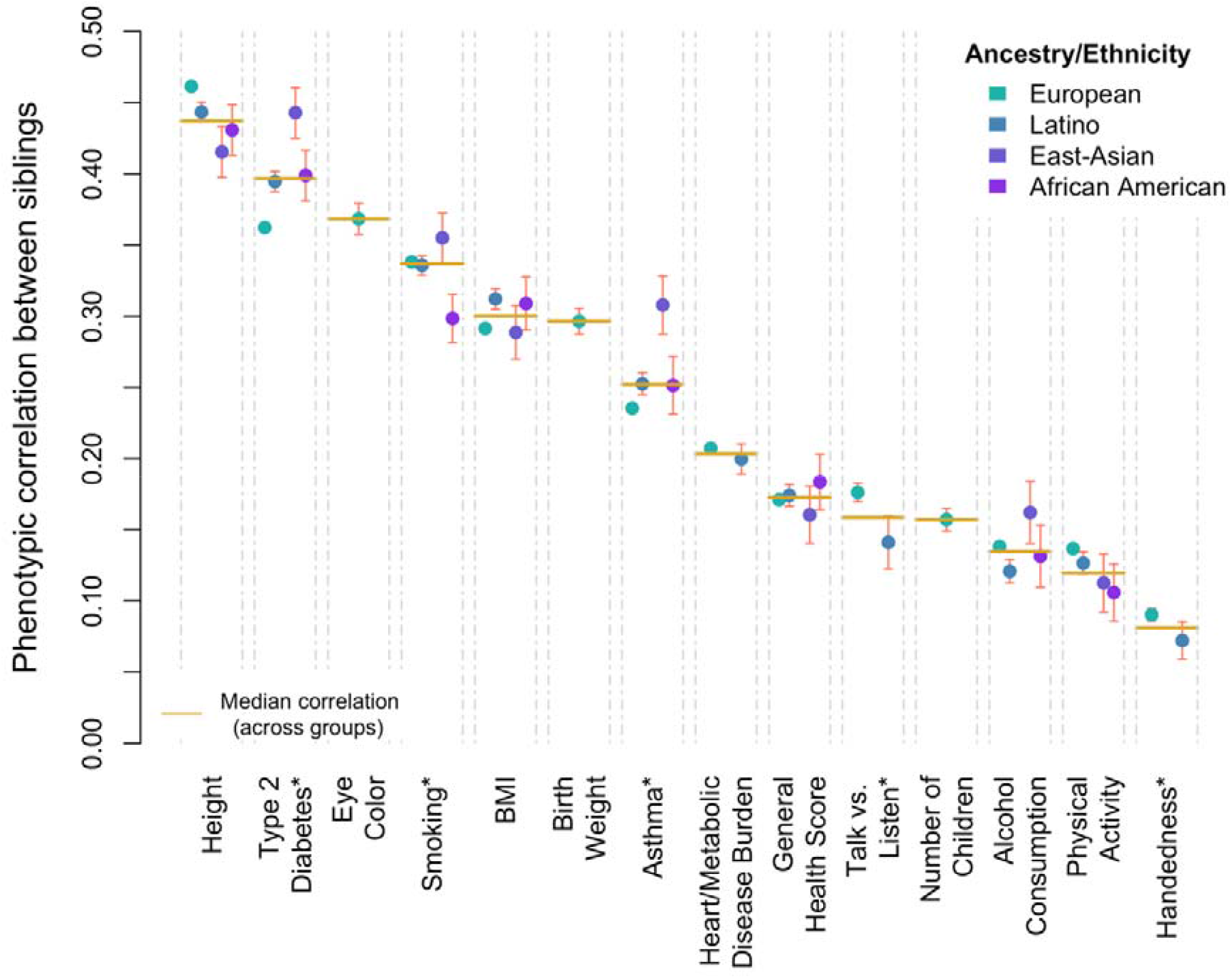
Estimates of phenotypic correlations between siblings across 14 phenotypes. Pearson correlation coefficients were calculated separately for each trait and ancestry/ethnicity group after adjusting phenotypes for age and sex. For binary traits (marked with a “*”), these estimates were further converted into tetrachoric correlations using an R function described in the METHODS section (Code Availability). Standard errors of sibling correlations () were approximated as 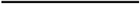, with being the corresponding number of pairs. Error bars represent 1.96 times the standard error. Numerical values and sample sizes are available in **Supplementary Table 4**.

### Validation using the model traits height and BMI

We first focus on the “model traits” height and BMI whose heritability had previously been estimated using a similar experimental design.^7–9^ Heritability estimates for height and BMI are shown in **Fig. 2a** and reported in **Table 1**, which also displays concordant phenotypic correlations for both traits across ancestry groups. For height, estimates of *h*^2^ ranged between 0.68 (standard error (s.e.) = 0.03) in the sub-cohort of EUR sibling pairs and 1.13 (s.e. 0.21) in siblings of East Asian ancestry (EAS). For BMI, 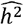 ranged between 0.47 (s.e. 0.04) in EUR and 0.68 (s.e. 0.24) in EAS siblings. We then performed an inverse-variance weighted meta-analysis of heritability estimates across the four sub-cohorts and obtained combined estimates for height and BMI of 0.70 (s.e. 0.03) and 0.48 (0.03), respectively (**Table 1**). Overall, we found no significant heterogeneity in heritability estimates across ancestry groups (Cochran’s heterogeneity test *P*_*HET*_ ≥ 0.2; **Table 2**). We also found no significant heterogeneity between estimates from this study and those from Sidorenko et al.^8^ (*P*_*HET*_ ≥ 0.2), although estimates from this study were a bit lower likely due to measurement errors in self-reported traits of 23andMe, Inc. participants. By combining our results with those of Sidorenko et al.^8^ and Young et al.^7^ we reached an unprecedented sample size of 680,242 sibling pairs for height and 671,856 sibling pairs for BMI and obtained meta-analysed FSIR estimates of 0.71 (s.e. 0.02) for height and 0.49 (s.e. 0.03) for BMI (**Table 1**). Importantly, FSIR is known to produce downwardly biased estimates of heritability for traits driving mate choice (for example, height), but these biases can be corrected if the mate correlation is known.^7^ Therefore, assuming a correlation of ∼0.2 between mates’ heights,^10^ our FSIR estimate of 0.71 (s.e. 0.02) can be converted into an estimated heritability of height of 0.86 (s.e. 0.03) in the current population (Methods).

**Figure 2.**
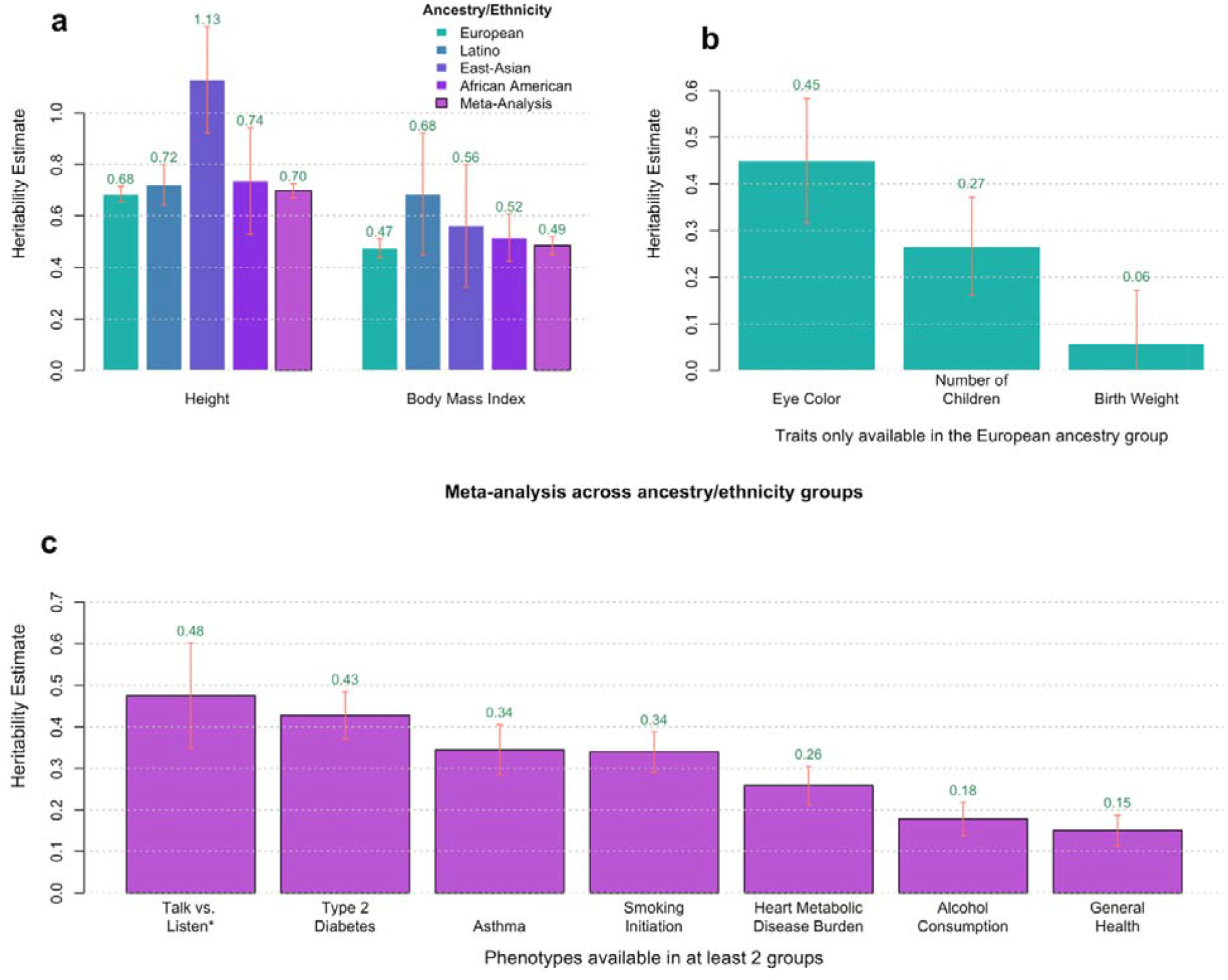
Estimates of heritability based on within-family Mendelian segregation in ∼500,000 sibling pairs. Estimates were obtained using restricted maximum likelihood in four ancestry/ethnicity group separately then meta-analysed across groups using the fixed-effect method. Panel **a** focuses on height and body mass index. Panel **b** focuses on traits only available in the European ancestry group. Panel c displays meta-analysed estimates for 7 traits present in a least two groups. Phenotypes marked with a star (*) were only reported by European ancestry and Latino ethnicity individuals. Each bar represents a point estimate, and the corresponding error bar represents its standard error (s.e.). Estimates for binary traits are reported on the liability scale (METHODS; **Supplementary Note 1**). Numerical values for panels **a** and **b** are given in **Table 1** and **Supplementary Table 4**, respectively, while those underlying panel **c** are available in **Table 2**. Prevalence of binary traits are reported for each sub-cohort in **Supplementary Table 2**. Estimated variance components were not constrained to be positive to ensure unbiasedness.

**Table 1.** FSIR estimates of heritability for height and body mass index. Estimates of heritability 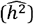 and residual correlation 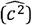 are reported for four sub-cohort within the 23andMe study, corresponding to four ancestry or ethnicity group. Phenotypic correlation between siblings were calculated after traits were adjusted for age and sex. Standard errors of sibling correlations (*r*_*s*_) were approximated as 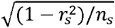, with *n*_*s*_ being the corresponding number of pairs. Data from Young et al.^7^ were obtained from a previous meta-analysis performed by Kemper and colleagues.^11^

| Trait | Cohort / Study | Number of sibling pairs | Sibling Correlation (s.e.) | $\widehat{h^2}$ (s.e.) | $\widehat{c^2}$ (s.e.) |
| --- | --- | --- | --- | --- | --- |
| Height | 23andMe (European) | 412,590 | 0.46 (0.001) | 0.68 (0.03) | 0.12 (0.01) |
|  | 23andMe (Latino) | 63,485 | 0.44 (0.003) | 0.72 (0.08) | 0.08 (0.04) |
|  | 23andMe (East Asian) | 9,990 | 0.41 (0.009) | 1.13 (0.21) | -0.15 (0.10) |
|  | 23andMe (African American) | 9,873 | 0.43 (0.009) | 0.74 (0.20) | 0.06 (0.10) |
|  | **Meta-analysis (23andMe)** | **495,938** | **0.45 (0.001)** | **0.70 (0.03)** | **0.11 (0.01)** |
|  | Sidorenko et al. (European) | 119,457 | 0.48 (0.002) | 0.76 (0.05) | 0.11 (0.03) |
|  | Young et al. (European) | 64,847 | 0.41 (0.01) | 0.68 (0.10) | 0.07 (0.05) |
|  | **Meta-analysis (23andMe + Young et al. + Sidorenko et al.)** | **680,241** | **0.46 (0.001)** | **0.71 (0.02)** | **0.10 (0.01)** |
| Body Mass Index | 23andMe (European) | 412,590 | 0.29 (0.001) | 0.47 (0.04) | 0.05 (0.02) |
|  | 23andMe (Latino) | 63,485 | 0.31 (0.003) | 0.51 (0.09) | 0.05 (0.05) |
|  | 23andMe (East Asian) | 9,990 | 0.29 (0.009) | 0.68 (0.24) | -0.06 (0.12) |
|  | 23andMe (African American) | 9,873 | 0.31 (0.009) | 0.56 (0.24) | 0.02 (0.12) |
|  | **Meta-analysis (23andMe)** | **495,938** | **0.29 (0.001)** | **0.49 (0.03)** | **0.05 (0.02)** |
|  | Sidorenko et al. (European) | 119,457 | 0.27 (0.002) | 0.55 (0.07) | -0.01 (0.04) |
|  | Young et al. (European) | 56,461 | 0.27 (0.01) | 0.39 (0.12) | 0.08 (0.06) |
|  | **Meta-analysis (23andMe + Young et al. + Sidorenko et al.)** | **671,856** | **0.29 (0.001)** | **0.49 (0.03)** | **0.04 (0.01)** |

### Behavioural traits

The results for height and BMI are consistent with previous estimates and therefore imply adequate quality of self-reported phenotypes for this experimental design. We therefore extended our analyses to additional phenotypes starting with four behavioural traits (**Fig. 2b**). Note that these were not available in all ancestries. We find substantial heritability for smoking initiation (0.34, s.e. 0.05), alcohol use (0.18, s.e. 0.04), number of children (meta-analysis across sexes: 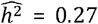, s.e. 0.10) and personality (“talk versus listen”: 0.48, s.e. 0.13). Sex-specific estimates for number of children are reported in **Supplementary Table 4**. Taken together, the results for these phenotypes imply that a significant proportion of phenotypic variation is due to direct genetic effects. These traits include substance use and reproductive fitness (number of children).

### Health and disease phenotypes

Many common diseases have known risk factors that are associated with behaviour. For example, a high-fat high-carb diet is a risk factor for type 2 diabetes, and smoking, pet ownership and diet affect risk of asthma. We focussed on health and disease associated phenotypes (**Fig. 2c**). For the common diseases type-2 diabetes and asthma, we find substantial heritability, 0.43 (s.e. 0.06) and 0.34 (s.e. 0.06), respectively, as well as for the self-reported burden of heart-metabolic disease (0.26, s.e. 0.05) and general health score (0.15, s.e. 0.04). To our knowledge, these are the first-ever within-family estimates of common diseases and health status. We also analysed self-reported physical activity, which had a lower estimate of 0.10 (s.e. 0.04; **Table 2**).

**Table 2.** Meta-analysed estimates of heritability for 11 traits available in at least two ancestry groups. Phenotypes marked with a star (*) were meta-analysed across European ancestry and Latino ethnicity study participants, while estimates for other traits were combined across all four groups. Standard errors (s.e.) of estimates of heritability 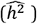 and residual correlation 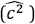 are reported between brackets under their corresponding estimate. Cochran heterogeneity p-value (*P*_*HET*_) are reported for each trait and focal parameter. Prevalence of binary traits are reported for each sub-cohort in **Supplementary Table 2**. P-values refer to two-sided Wald-test.

| Trait/Disease | Family of Traits | Trait Distribution | Number of sibling pairs | $\widehat{h}^2$<br>(s.e.) | P-value for $\widehat{h}^2$ | $P_{HET}$ for $\widehat{h}^2$ | $\widehat{c}^2$<br>(s.e.) | P-value for $\widehat{c}^2$ | $P_{HET}$ for $\widehat{c}^2$ |
| --- | --- | --- | --- | --- | --- | --- | --- | --- | --- |
| Height | Model Trait | Quantitative | 495,938 | 0.70<br>(0.03) | $1.5 \times 10^{-141}$ | 0.20 | 0.11<br>(0.01) | $3.8 \times 10^{-14}$ | 0.07 |
| Body Mass Index | Model Trait | Quantitative | 495,938 | 0.49<br>(0.03) | $4.8 \times 10^{-47}$ | 0.80 | 0.05<br>(0.02) | $3.9 \times 10^{-3}$ | 0.83 |
| General Health Score | Disease and Health | Quantitative | 487,916 | 0.15<br>(0.04) | $5.1 \times 10^{-5}$ | 0.24 | 0.10<br>(0.02) | $3.6 \times 10^{-7}$ | 0.23 |
| Heart/Metabolic Disease Burden* | Disease and Health | Quantitative | 296,356 | 0.26<br>(0.05) | $3.0 \times 10^{-8}$ | 0.13 | 0.08<br>(0.02) | $1.3 \times 10^{-3}$ | 0.11 |
| Type 2 Diabetes | Disease and Health | Binary | 507,220 | 0.43<br>(0.06) | $5.3 \times 10^{-14}$ | 0.03 | 0.25<br>(0.05) | $1.9 \times 10^{-6}$ | $8.90 \times 10^{-4}$ |
| Asthma | Disease and Health | Binary | 459,461 | 0.34<br>(0.06) | $1.2 \times 10^{-8}$ | 0.45 | 0.07<br>(0.04) | 0.07 | 0.84 |
| Smoking Initiation | Behavioural | Binary | 547,126 | 0.34<br>(0.05) | $1.2 \times 10^{-12}$ | 0.46 | 0.17<br>(0.03) | $1.5 \times 10^{-9}$ | 0.38 |
| Talk versus Listen* | Behavioural | Binary | 98,725 | 0.48<br>(0.13) | $1.7 \times 10^{-4}$ | 0.87 | -0.07<br>(0.07) | 0.35 | 1.00 |
| Alcohol Consumption | Behavioural | Quantitative | 428,371 | 0.18<br>(0.04) | $1.1 \times 10^{-5}$ | 0.73 | 0.05<br>(0.02) | 0.02 | 0.87 |
| Handedness* | Behavioural | Binary | 195,307 | 0.16<br>(0.14) | 0.24 | 0.81 | 0.01<br>(0.09) | 0.93 | 0.88 |
| Physical Activity | Behavioural | Quantitative | 470,327 | 0.10<br>(0.04) | 0.01 | 0.20 | 0.09<br>(0.02) | $1.1 \times 10^{-5}$ | 0.33 |

### Lack of heterogeneity across ancestries

Overall, eleven traits were measured in at least two sub-cohorts. Among those (and excluding height and BMI already discussed above), we found significant heritability for 7 traits (**Table 2**). Heritability estimates for these 7 traits were all significant in the EUR sub-cohort, while only 2 out of 7 reached statistical significance in the sub-cohort of siblings of Latino ethnicity (Supplementary Table 4). Furthermore, we meta-analysed heritability across ancestry groups. Like height and BMI, we found no significant heterogeneity in heritability estimates across ancestry groups (*P*_*HET*_ > 0.1) for all phenotypes except type 2 diabetes (T2D: *P*_*HET*_ = 0.03; **Table 2**). Importantly, a lack of significant heterogeneity may also reflect low statistical power in under-sampled ancestry groups from this study. After meta-analysis across sub-cohorts, we did not detect significant heritability estimates for more phenotypes than the 7 traits already highlighted above. Meta-analysed estimates for all 11 phenotypes are reported in **Table 2**.

### Common environmental effects for behaviour and disease phenotypes

The FSIR method also provides estimates of the proportion of trait variance (hereafter denoted *c*^2^, and its estimate 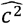) due to phenotypic effects that are common to siblings yet independent of IBD sharing. Note that a positive ^2^ is also expected under assortative mating,^11^ and that unmodelled non-additive genetic effects contribute negative *c*^2^ (**Supplementary Note 2**). Therefore, this component could contain both genetic and shared environmental variance. Estimates of *c*^2^ for all ancestry groups and traits are reported in **Supplementary Table 4**. Among traits with significant heritability estimates, we found a significant 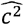 only in the EUR sub-cohort for smoking initiation (-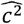 = 0.17, s.e. 0.03), height (--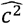 = 0.12, s.e. 0.01), heart or metabolic disease burden (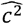= 0.09, s.e. 0.02) and general health score (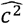 = 0.08, s.e 0.02). We also find a significant 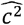 in the EUR sub-cohort for birth weight (BW: 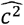= 0.27, s.e 0.06) and physical activity (PA: 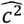 = 0.07, s.e. 0.02), for which the corresponding heritability estimates did not reach statistical significance (BW: 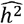= 0.06, s.e. 0.11; PA: 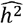= 0.13, s.e. 0.04). Finally, we detected a significant 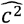 for type 2 diabetes (T2D) after meta-analysing estimates across ancestry groups (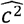= 0.25, s.e. 0.05) with a significant heterogeneity between groups (**Table 2**).

We compared our within-family estimates with between-family estimates from previous heritability studies of T2D and asthma,^12^ smoking initiation,^13^ height and BMI,^11^ and number of children.^14^ We focused on these six traits and studies because the between-family estimates were precise enough (defined here by s.e. lower than 0.05) to ensure a meaningful comparison. Overall, we observed remarkably similar heritability estimates across the two designs (**Table 3**), which suggests that assumptions underlying between-family studies (for example, the assumption that shared environmental effects are constant for different types of relatives) may hold true for most traits. However, there was a notable difference for T2D for which Speed and Evans^12^ reported a liability-scale estimate of 0.62 (s.e 0.04), consistent with previous between-family estimates from the literature,^15^ yet significantly larger than our estimate of 0.43 (s.e. 0.06; **Table 2**). Given the substantial 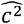 reported above for T2D (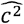 = 0.25, s.e 0.05; **Table 2**), our data suggest that the difference between 0.62 and 0.43 may be caused by shared environmental effects or unaccounted population stratification, which might have inflated between-family estimates.

**Table 3.**
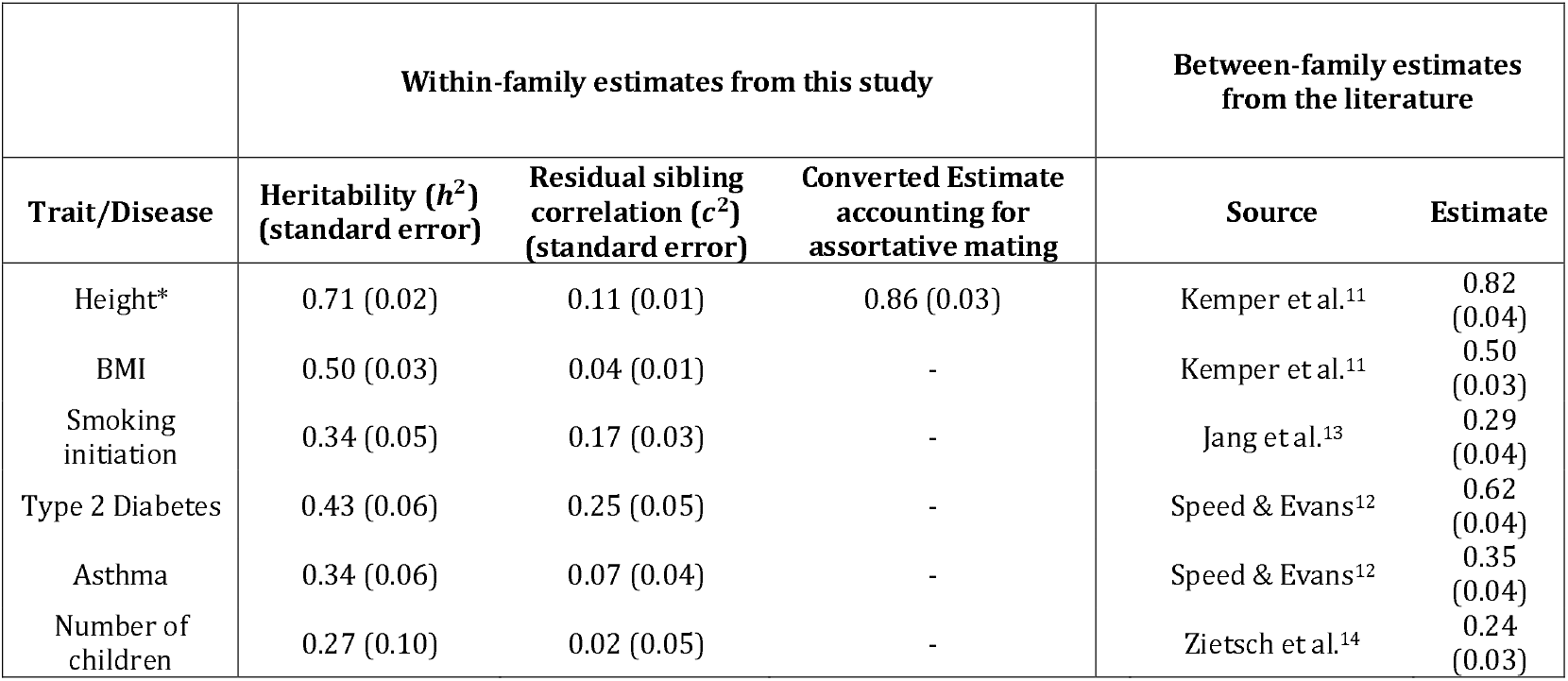
Comparison of within-family versus between-family heritability estimates across six phenotypes. *For eight, within-family estimates were converted into estimates of heritability in the current population by assuming a pousal correlation *r* = 0.2.

|  | Within-family estimates from this study |  |  | Between-family estimates from the literature |  |
| --- | --- | --- | --- | --- | --- |
| Trait/Disease | Heritability ($h^2$)<br>(standard error) | Residual sibling correlation ($c^2$)<br>(standard error) | Converted Estimate accounting for assortative mating | Source | Estimate |
| Height* | 0.71 (0.02) | 0.11 (0.01) | 0.86 (0.03) | Kemper et al.<sup>11</sup> | 0.82 (0.04) |
| BMI | 0.50 (0.03) | 0.04 (0.01) | - | Kemper et al.<sup>11</sup> | 0.50 (0.03) |
| Smoking initiation | 0.34 (0.05) | 0.17 (0.03) | - | Jang et al.<sup>13</sup> | 0.29 (0.04) |
| Type 2 Diabetes | 0.43 (0.06) | 0.25 (0.05) | - | Speed & Evans<sup>12</sup> | 0.62 (0.04) |
| Asthma | 0.34 (0.06) | 0.07 (0.04) | - | Speed & Evans<sup>12</sup> | 0.35 (0.04) |
| Number of children | 0.27 (0.10) | 0.02 (0.05) | - | Zietsch et al.<sup>14</sup> | 0.24 (0.03) |

## DISCUSSION

We have used the largest sample size (number of sibling pairs) to date to study behavioural and behaviour-associated health and disease traits by contrasting the genome-wide realised relationship between siblings with their phenotypic similarity. We report robust FSIR heritability estimates for 12 complex traits, including multiple behavioural phenotypes and 2 common diseases in a large sample of genetically diverse sibling pairs. Standard errors of our FSIR heritability estimates are lower than 0.1 for 9 phenotypes, thus reaching unprecedented precision for most traits investigated using this experimental design. Overall, our results show a substantial amount of genetic variation underlying behavioural and anthropometric traits, as well as susceptibility to type 2 diabetes and asthma, a fraction of which has yet to be accounted for by GWAS to date.^16^

Our results show a remarkable concordance of sibling correlations (**Table 1**) and estimates of heritability across ancestry groups. This result is not a foregone conclusion because different ancestry groups can differ in both their environment and in the amount of genetic variation. For example, there is more genetic variation among populations of African ancestry^17^ and therefore, everything else being the same one may have expected a larger heritability for such populations. However, differences between ancestry-divergent populations in genomic variation are small and we do not have enough statistical power to detect the effect of such small differences on heritability estimates. In addition, natural selection has partially shaped trait-associated variation, and we do not have sufficient information on past selection to predict how genetic variation is shaped between and within populations.

The estimates from within-family segregation variance are generally much closer to estimates from pedigree or twin designs (**Table 3**) than estimates captured from GWAS data (SNP-heritability), even when the latter is from whole-genome sequence data. For example, the estimates of heritability for BMI from within-family and pedigree designs are approximately 0.5 (**Table 3**) yet estimates from GWAS data are approximately 0.3 (ref.^18,19^). It is not clear if the discrepancy if because population-based estimates are biased downward due to gene-environment interactions or if the within-family estimates are biased upwards due to non-additive genetic variation. It is in theory possible to separate additive from non-additive genetic variation using the within-family design, but much larger sample sizes (of the order of millions of sibling pairs) would be required to separate such effects.^8^

There is no consensus in the literature on the magnitude of additive genetic variation associated with cognitive ability and educational attainment (EA). Reported within-family GWAS estimates (analogous to SNP-heritability) for EA are very low (0.04, ref.^20^), but pedigree-based (0.43 s.e. 0.04) and FSIR estimates from Young et al.^7^ in Iceland 0.40 (s.e. 0.15) were much larger. Moreover, the estimates from within-family GWAS and within-family IBD are biased downwards because of assortative mating.^8,11^ We could not analyse EA and cognitive traits because of ethics restrictions. However, we emphasise that additional research using the within-family is needed to inform ongoing debates about the relative importance of direct and indirect genetic effects for such traits.

## CONSORTIUM MEMBERS

We list below members of the 23andMe Research team (for data generated in 2024) and highlight in bold font those listed as authors.

**23andMe Research Team** (contributing authors on this manuscript are highlighted in bold font) Stella Aslibekyan, Adam Auton, Elizabeth Babalola, Robert K. Bell, Jessica Bielenberg, Ninad S. Chaudhary, Zayn Cochinwala, Sayantan Das, Emily DelloRusso, Payam Dibaeinia, Sarah L. Elson, Nicholas Eriksson, Chris Eijsbouts, Teresa Filshtein, Pierre Fontanillas, Davide Foletti, Will Freyman, Zach Fuller, **Julie M. Granka**, Chris German, Éadaoin Harney, Alejandro Hernandez, Barry Hicks, David A. Hinds, M. Reza Jabalameli, Ethan M. Jewett, Yunxuan Jiang, Sotiris Karagounis, Lucy Kaufmann, Matt Kmiecik, Katelyn Kukar, Alan Kwong, Keng-Han Lin, **Yanyu Liang**, Bianca A. Llamas, Aly Khan, Steven J. Micheletti, Matthew H. McIntyre, Meghan E. Moreno, Priyanka Nandakumar, Dominique T. Nguyen, Jared O’Connell, Steve Pitts, G. David Poznik, Alexandra Reynoso, Shubham Saini, Morgan Schumacher, Leah Selcer, Anjali J. Shastri, Jingchunzi Shi, Suyash Shringarpure, Keaton Stagaman, Teague Sterling, Qiaojuan Jane Su, Joyce Y. Tung, Susana A. Tat, Vinh Tran, **Xin Wang**, Wei Wang, Catherine H. Weldon, Amy L. Williams, Peter Wilton.

## Acknowledgements

We acknowledge the research participants and employees of 23andMe for making this work possible. L.Y. is supported by the Australian Research Council (FT220100069) and the Snow Medical Research Foundation. P.M.V. was supported by the Australian Research Council (FL180100072). D.M.E. is supported by an Australian National Health and Medical Research Council Investigator Award (2017942).

## Author contributions

P.M.V. and L.Y. conceived and jointly supervised the study.

Y.L., X.W. and J.M.G. conducted statistical analyses of 23andMe Inc. data with the assistance or guidance from J.S., D.M.E., P.M.V., and L.Y.

L.Y., P.M.V., J.S. and D.M.E. wrote the manuscript with the participation of all authors. All the authors approved the final version of the manuscript.

## Competing interests

Y.L. and J.M.G. are employed by and hold stock or stock options in 23andMe, Inc. X.W. is a former employee of 23andMe Inc. Other authors have no competing interests to declare.

## METHODS

### Ethics declaration

Individuals included in this study are research participants of 23andMe, Inc., a consumer genetics and research company who were genotyped as part of the 23andMe Personal Genome Service. 23andMe participants provided informed consent and volunteered to participate in the research online, under a protocol approved by the external AAHRPP-accredited IRB, Ethical & Independent (E&I) Review Services. As of 2022, E&I Review Services is part of Salus IRB (https://www.versiticlinicaltrials.org/salusirb). In the current study, we used research-consented individuals providing self-reported phenotypes, with at least one genetically inferred full sibling also in the cohort. The total sample size of 23andMe participants analysed in the current study is 1,024,523.

### Genotyping and Phenotyping

Sample and trait selection. Ancestry (or ethnicity) groups were determined using 23andMe, Inc.’s genetic ancestry classifier,^21^ which assigns the following ancestry/ethnicity labels: African American (AFR), European (EUR), East Asian (EAS), and Latino (LAT). We focused our analyses on EUR, EAS, AFR and LAT full sibling pairs with available (self-reported) phenotype measurements for a minimum of 5,000 pairs per trait-ancestry combination. We also focused on diseases with a prevalence larger than 3%. Altogether, 14 traits were selected for analysis (**Supplementary Table 1**). Note that number of children was recorded (and primarily analysed) as separate traits for male and female participants.

### Sibship inference

We started with 10,317,496 23andMe, Inc. participants who had consented to research and were classified as the four genetic ancestry/ethnicity groups. Among those, a sibling relation between two individuals was inferred if the lengths (in centimorgan units; cM) of genomic segments with exactly one IBD haplotype IBD ranged between 2,249 cM and 3,373 cM, and that of segments where both haplotypes are IBD ranged between 375 cM and 2,249 cM, as previously described.^22^ FSIR analyses were based on quasi-independent sib-pairs, a simplified data structure assuming sibling pairs to be independent even when the siblings involved were from the same family. For example, a sibship of four siblings would lead to 4 × 3/2 = 6 quasi-independent sib-pairs included in our analysis.

### SNP selection for IBD inference

Genotyping was performed on several arrays. Approximately 25,000 to 50, 000 directly genotyped and LD-pruned SNPs were used for IBD inference. Specific numbers of SNPs are reported in **Supplementary Table 3**. LD-pruning was performed within each sub-group. We selected SNPs that were available on all genotyping arrays, and which had a MAF >5% (in the corresponding ancestry group), a missing genotype rate <1%, a *p*-value for Hardy-Weinberg test larger than 10^−8^, and a pair-wise LD r^2^ below 0.1 between SNPs less than 1Mb apart. Genomic positions used in this study refers to the hg38 genome build. We used the sex-averaged genetic map from Haldorsson et al.^23^ to convert genomic positions into genetic distances.

Phenotype quality control. We performed phenotype adjustments within ancestry groups and separately for males and females. Outlier phenotype values (> 6 SD away from the mean) were excluded. Phenotypes were then residualized after fitting age (at assessment) and its square value as covariates in a linear regression model (for binary phenotype, cases were treated as 1 and controls were treated as 0). The resulting residuals were further scaled to have a mean of 0 and a variance of 1 within each sex. Ancestry/ethnicity group specific phenotypic means, SDs, and age distributions are provided for males and females separately in **Supplementary Table 1**. Covariate-adjusted phenotypic correlations are reported in **Supplementary Table 4**. For binary traits, these adjusted correlations were converted into tetrachoric correlations using a custom R function (see Code Availability).

### IBD estimation and heritability estimation

#### Calculation of IBD coefficients

The mean and standard deviation of IBD coefficients for each ancestry sub-group is presented in **Supplementary Table 3**. IBD coefficients between siblings were calculated using the lengths of genomic segments shared IBD, inferred using the ibd.py module of the *snipar* software tool (Code Availability).^24^ Each IBD segment was labelled as IBD1 or IBD2 if 1 or the 2 haplotypes forming the segment was IBD, respectively. IBD segments were called assuming a genotyping error rate of 0.00045.^25^ This parameter was set using the -- p_error flag in *snipar*. We calculated a genome-wide IBD coefficient for each sibling pair as *L*_1_/2 + *L*_2_ /*L*_*G*_, where *L*_1_ and *L*_2_ are the cumulated lengths (in centimorgan units) of IBD1 and IBD2 segments, respectively, and *L*_*G*_ is the total length of the genome.

#### Recombination rate stratified IBD coefficients

First, we stratified 1,122,949 genomic segments forming the (sex-averaged) recombination map from Haldorsson et al.^23^ into recombination rates (RR) quartile groups. Next, we calculated a genome-wide IBD coefficient for each RR quartile group using the cumulative lengths of the IBD segments within each group as described above.

#### Heritability estimation

We estimated *h*^2^ and *c*^2^ using the REstricted Maximum Likelihood (REML) implemented in a custom R-script available on Zenodo (see Code Availability).^26^ Estimates for binary traits were obtained on an observed 0-1 scale then converted into a Gaussian liability scale using the formalism described in **Supplementary Note 1**. We provide an R script to convert estimates from the observed to the liability scale, which implements the method described in **Supplementary Note 1** (see Code Availability). For RR-stratified analyses, we jointly fitted IBD coefficients for each RR group into our heritability model and obtained our final estimate by summing the contributions across all groups as previously described.^8^ As in previous studies,^8,11^ we used Equation (2) combined with estimates of the correlation between mates’ phenotypes 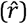 to convert 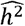 into an estimate 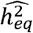 of heritability in a population undergoing assortative mating, and also to derive its standard error:

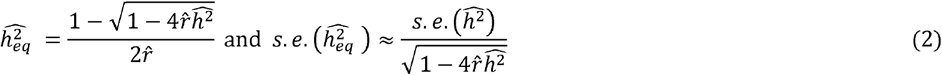

## DATA AVAILABILITY

Application for data access can be submitted at https://research.23andme.com/dataset-access/.

## CODE AVAILABILITY

Statistical analyses were performed using R (v4.1.0, 4.2.1, https://cran.r-project.org/).

The R function used to estimate heritability using restricted maximum likelihood estimation is available on Zenodo.^26^

The R functions used to convert estimates for binary traits from the observed 0-1 scale to an underlying Gaussian scale, and Pearson correlation coefficients to tetrachoric correlation coefficients are available on GitHub here: https://github.com/loic-yengo/ConvertFSIR_to_liability

Snipar software was used to estimate IBD sharing: https://snipar.readthedocs.io/en/latest/guide.html

Genotype data quality control, including filtering and LD pruning, as well as allelic scoring was performed with PLINK v2.00a5.9LM (https://www.cog-genomics.org/plink/2.0/) and PLINK v1.90b6.25 (https://www.cog-genomics.org/plink/).

