## Supplementary Figures and Notes for "Within-family heritability estimates for behavioural and disease phenotypes from 500,000 sibling pairs of diverse ancestries"

**Supplementary Note 1: Converting estimates from the observed 0-1 scale to an underlying normally distributed liability scale**

### *1.1. Notations and definitions*

Let us consider a pair of siblings $i$ and $j$. We denote $\ell_i$ and $\ell_j$ as their underlying continuous liability values and $y_i$ and $y_j$ as their binary disease phenotype, respectively. Our derivation presented below assumes a liability threshold model (Falconer 1965) with $\ell_i$ following a Gaussian distribution with mean 0 and variance 1. Under this model the relationship between $\ell_i$ and $y_i$ only depends on the disease prevalence $(K)$ and can be expressed as $y_i = \mathbb{I}_{\ell_i > t}$, where $\mathbb{I}_{\ell_i > t}$ is the indicator that $\ell_i$ exceeds $t = \Phi^{-1}(1-K)$ with $\Phi$ being the cumulative density function of a standard normal distribution. We also denote $\phi(t)$ as the probability density function of the standard normal distribution evaluated at the threshold $t$.
We further assume that all resemblance between relatives (on the liability scale) is due to additive genetic effects and other factors shared between siblings are uncorrelated with the realised relationship coefficient between siblings, hereafter denoted $G_{ij}$. We assume that the prevalence is known as well as the distribution of $G_{ij} \sim N(0.5, s^2)$, where $s \approx 0.038$ (Visscher et al. 2006).[3]
Therefore, we can express the covariance between $\ell_i$ and $\ell_j$ as

$$\text{(1.1)} \qquad \mathrm{cov}(\ell_i, \ell_j) = c_\ell^2 + h_\ell^2 G_{ij},$$

where $h_\ell^2$ denotes the heritability on the liability scale and $c_\ell^2$ denotes the proportion of liability variance unexplained by additive genetic effects. For example, $c_\ell^2$ can be caused by environmental effects shared by siblings.

We define the heritability on the observed 0-1 scale as

$$\text{(1.2)} \qquad h_{01}^2 = \frac{\mathrm{cov}[(y_i - K)(y_j - K), G_{ij}]}{K(1-K)\mathrm{var}(G_{ij})} = \frac{\mathrm{E}[(y_i - K)(y_j - K)G_{ij}] - \mathrm{E}[(y_i - K)(y_j - K)]\mathrm{E}[G_{ij}]}{K(1-K)\mathrm{var}(G_{ij})}$$

and the residual correlation between siblings $c_{01}^2$ as

$$\text{(1.3)} \qquad c_{01}^2 = \frac{\mathrm{E}[(y_i - K)(y_j - K)]}{K(1-K)} - \frac{h_{01}^2}{2} = r_{01} - \frac{h_{01}^2}{2}$$

such that $r_{01} = \mathrm{E}[(y_i - K)(y_j - K)]/[K(1-K)]$ is the sibling correlation on the observed 0-1 scale, which can also be written as

$$\text{(1.4)} \qquad r_{01} = \frac{\mathrm{E}[y_i y_j] - K^2}{K(1-K)}$$

Moreover, $\mathrm{E}[y_i y_j]$ can be expressed as

$$\text{(1.5)} \qquad \mathrm{E}[y_i y_j] = \mathrm{E}\left[\mathrm{E}[y_i y_j | G_{ij}]\right] = I_0(c_\ell^2, h_\ell^2, t) = \int F(c_\ell^2 + h_\ell^2 x; t)\, \phi\left(\frac{x - 0.5}{s}\right)/s\, dx,$$

where $F(c_\ell^2 + h_\ell^2 x; t)$ denotes the top-right quadrant probability above thresholds $(t, t)$ of a standard bivariate Gaussian distribution with correlation $c_\ell^2 + h_\ell^2 x$. Note that $F(c_\ell^2 + h_\ell^2 x; t)$ does not have a closed-form expression but can be calculated numerically using R packages such as `fMultivar` or `mvtnorm`.

### *1.2. Relationship between $(c_\ell^2, h_\ell^2)$ and $(c_{01}^2, h_{01}^2)$*

Equation (1.2) can be rewritten as

(1.6) $h_{01}^2 = \frac{\mathrm{E}[(y_i-K)(y_j-K)G_{ij}]}{K(1-K)\mathrm{s}^2} - \frac{r_{01}}{2s^2}.$

We can calculate $\mathrm{E}[(y_i - K)(y_j - K)G_{ij}]$ as:

$$
\begin{aligned}
(1.7)\quad \mathrm{E}[(y_i - K)(y_j - K)G_{ij}] &= \mathrm{E}[y_i y_j G_{ij}] + K^2 E[G_{ij}] - K\left[\mathrm{E}[y_i G_{ij}] + \mathrm{E}[y_j G_{ij}]\right] \\
&= \mathrm{E}[y_i y_j G_{ij}] + K^2 E[G_{ij}] - 2K\mathrm{E}[y_i]\mathrm{E}[G_{ij}] \\
&= \mathrm{E}[y_i y_j G_{ij}] + K^2 E[G_{ij}] - 2K^2 E[G_{ij}] \\
&= \mathrm{E}[y_i y_j G_{ij}] - K^2 E[G_{ij}] = \mathrm{E}[y_i y_j G_{ij}] - K^2/2
\end{aligned}
$$

Furthermore, $\mathrm{E}[y_i y_j G_{ij}]$ can be expressed as

$$(1.8)\quad \mathrm{E}[y_i y_j G_{ij}] = \mathrm{E}[\mathrm{E}[y_i y_j | G_{ij}]G_{ij}] = I_1(c_\ell^2, h_\ell^2, t) = \int xF(c_\ell^2 + h_\ell^2 x; t)\,\phi\left(\frac{x-0.5}{s}\right)/s\,dx$$

Therefore,

$$(1.9)\quad \mathrm{E}[(y_i - K)(y_j - K)G_{ij}] = I_1(c_\ell^2, h_\ell^2, t) - K^2/2$$

Similarly,

$$(1.10)\quad \mathrm{E}[(y_i - K)(y_j - K)] = \mathrm{E}[y_i y_j] - K^2 = I_0(c_\ell^2, h_\ell^2, t) - K^2$$

which leads to

$$
\begin{aligned}
(1.11)\quad h_{01}^2 &= \frac{I_1(c_\ell^2, h_\ell^2, t) - K^2/2 - I_0(c_\ell^2, h_\ell^2, t)/2 + K^2/2}{K(1-K)\mathrm{s}^2} \\
&= \frac{I_1(c_\ell^2, h_\ell^2, t) - I_0(c_\ell^2, h_\ell^2, t)/2}{K(1-K)\mathrm{s}^2}
\end{aligned}
$$

Combining Eqns. (1.3) and (1.10) leads to

$$
\begin{aligned}
(1.12)\quad c_{01}^2 &= \frac{I_0(c_\ell^2, h_\ell^2, t) - K^2}{K(1-K)} - \frac{h_{01}^2}{2} \\
&= \frac{I_0(c_\ell^2, h_\ell^2, t) - K^2}{K(1-K)} - \left(\frac{I_1(c_\ell^2, h_\ell^2, t)/2 - I_0(c_\ell^2, h_\ell^2, t)/4}{K(1-K)\mathrm{s}^2}\right) \\
&= \frac{s^2 I_0(c_\ell^2, h_\ell^2, t) - s^2K^2 - I_1(c_\ell^2, h_\ell^2, t)/2 + I_0(c_\ell^2, h_\ell^2, t)/4}{K(1-K)\mathrm{s}^2} \\
&= \frac{\left(\frac{1}{4} + s^2\right) I_0(c_\ell^2, h_\ell^2, t) - I_1(c_\ell^2, h_\ell^2, t)/2 - s^2K^2}{K(1-K)\mathrm{s}^2}
\end{aligned}
$$

Altogether, the relationship between $(c_\ell^2, h_\ell^2)$ and $(c_{01}^2, h_{01}^2)$ can be summarized using the following set of equations:

$$(1.13)\quad \begin{cases} c_{01}^2 = \dfrac{\left(\frac{1}{4} + s^2\right) I_0(c_\ell^2, h_\ell^2, t) - I_1(c_\ell^2, h_\ell^2, t)/2 - s^2K^2}{K(1-K)\mathrm{s}^2} \\ h_{01}^2 = \dfrac{-I_0(c_\ell^2, h_\ell^2, t)/2 + I_1(c_\ell^2, h_\ell^2, t)}{K(1-K)\mathrm{s}^2} \end{cases} \Leftrightarrow (c_{01}^2, h_{01}^2) = \boldsymbol{H}[(c_\ell^2, h_\ell^2)]$$

where $\boldsymbol{H}$ is the bivariate function to conversion parameters between scales. We assume that $\boldsymbol{H}$ is bijective and can be inverted such that $(c_\ell^2, h_\ell^2) = \boldsymbol{H}^{-1}[(c_{01}^2, h_{01}^2)]$. In practice, we propose solving the following minimization problem to convert estimates on the observed 0-1 scale, hereafter denoted $(\widehat{c_{01}^2}, \widehat{h_{01}^2})$, into estimates on the liability scale, hereafter denoted $(\widehat{c_\ell^2}, \widehat{h_\ell^2})$:

$$(1.14) \qquad (\widehat{c_\ell^2}, \widehat{h_\ell^2}) = \boldsymbol{H}^{-1}[(\widehat{c_{01}^2}, \widehat{h_{01}^2})] \equiv \underset{(c_\ell^2, h_\ell^2)}{\operatorname{argmin}} \left\| \boldsymbol{H}[(c_\ell^2, h_\ell^2)] - (\widehat{c_{01}^2}, \widehat{h_{01}^2}) \right\|_2$$

We used the `optim` function of R to solve this problem. Note solutions to Eqn. (1.14) are unconstrained such that $\widehat{c_\ell^2}$ and $\widehat{h_\ell^2}$ could be outside of [0,1]. **Fig. S1-4** illustrate the performances of Eqn. (1.13) and (1.14) across various simulation scenarios.

### *1.3. Sampling variance for converted estimates of $(c_\ell^2, h_\ell^2)$*

We now assume that we have obtained an estimate of $(\widehat{c_{01}^2}, \widehat{h_{01}^2})$ with its sampling covariance matrix hereafter denoted $\Sigma_{01}$. We propose to approximate the sampling variance of converted estimate $(\widehat{c_\ell^2}, \widehat{h_\ell^2}) = \boldsymbol{H}^{-1}[(\widehat{c_{01}^2}, \widehat{h_{01}^2})]$ using the delta-method as

$$(1.15) \qquad \operatorname{var}[(\widehat{c_\ell^2}, \widehat{h_\ell^2})] = J[\boldsymbol{H}^{-1}[(\widehat{c_{01}^2}, \widehat{h_{01}^2})]]\, \Sigma_{01}\, J[\boldsymbol{H}^{-1}[(\widehat{c_{01}^2}, \widehat{h_{01}^2})]]$$

where $J[\boldsymbol{H}^{-1}[(\widehat{c_{01}^2}, \widehat{h_{01}^2})]]$ is the Jacobian of $\boldsymbol{H}^{-1}$ at $(\widehat{c_{01}^2}, \widehat{h_{01}^2})$, which can also be expressed as the inverse of the Jacobian of $\boldsymbol{H}$ at $(\widehat{c_\ell^2}, \widehat{h_\ell^2})$, i.e.

$$(1.16) \qquad J_{\boldsymbol{H}^{-1}[(\widehat{c_{01}^2}, \widehat{h_{01}^2})]} = \left(J_{\boldsymbol{H}[(\widehat{c_\ell^2}, \widehat{h_\ell^2})]}\right)^{-1}.$$

To derive the Jacobian of $\boldsymbol{H}$, we need partial derivatives of $I_0(c_\ell^2, h_\ell^2, t)$ and $I_1(c_\ell^2, h_\ell^2, t)$, with respect to $c_\ell^2$ and $h_\ell^2$. More specifically, $\boldsymbol{H}$ can be written in matrix form as

$$(1.17) \qquad \boldsymbol{H}\begin{pmatrix} c_\ell^2 \\ h_\ell^2 \end{pmatrix} = \begin{pmatrix} c_{01}^2 \\ h_{01}^2 \end{pmatrix} = \frac{1}{K(1-K)s^2}\begin{pmatrix} \frac{1}{4} + s^2 & -1/2 \\ -1/2 & 1 \end{pmatrix}\begin{bmatrix} I_0(c_\ell^2, h_\ell^2, t) \\ I_1(c_\ell^2, h_\ell^2, t) \end{bmatrix} + \frac{1}{K(1-K)s^2}\begin{pmatrix} -s^2K^2 \\ 0 \end{pmatrix}$$
$$= \frac{1}{K(1-K)s^2}\begin{pmatrix} \frac{1}{4} + s^2 & -1/2 \\ -1/2 & 1 \end{pmatrix}\begin{bmatrix} I_0(c_\ell^2, h_\ell^2, t) \\ I_1(c_\ell^2, h_\ell^2, t) \end{bmatrix} - \frac{1}{1-K}\begin{pmatrix} K \\ 0 \end{pmatrix}$$

which implies that

$$(1.18) \qquad J_{\boldsymbol{H}\begin{pmatrix} c_\ell^2 \\ h_\ell^2 \end{pmatrix}} = \frac{1}{K(1-K)s^2}\begin{pmatrix} \frac{1}{4} + s^2 & -1/2 \\ -1/2 & 1 \end{pmatrix} \times J\begin{bmatrix} I_0(c_\ell^2, h_\ell^2, t) \\ I_1(c_\ell^2, h_\ell^2, t) \end{bmatrix}$$

with

$$(1.19) \qquad J_{\begin{bmatrix} I_0(c_\ell^2, h_\ell^2, t) \\ I_1(c_\ell^2, h_\ell^2, t) \end{bmatrix}} = \begin{pmatrix} \dfrac{\partial I_0(c_\ell^2, h_\ell^2, t)}{\partial c_\ell^2} & \dfrac{\partial I_0(c_\ell^2, h_\ell^2, t)}{\partial h_\ell^2} \\ \dfrac{\partial I_1(c_\ell^2, h_\ell^2, t)}{\partial c_\ell^2} & \dfrac{\partial I_1(c_\ell^2, h_\ell^2, t)}{\partial h_\ell^2} \end{pmatrix}$$

In practice, we propose to calculate these derivatives numerically using the R package `numDeriv`.

**Supplementary Note 2: Effect of additive-by-additive interactions on FSIR estimates**

We use similar notations as in **Supplementary Note 1** and now assume that the phenotypic correlation between siblings $i$ and $j$ is partly caused by pairwise interactions between additive genetic effects across the genome. For simplicity, we also assume that phenotypes of siblings $i$ and $j$ (hereafter denoted $y_i$ and $y_j$, respectively) have a mean of 0 and a variance of 1.

Therefore,

(2.1) $$\mathrm{E}(y_i y_j | G_{ij}) = c_t^2 + h_t^2 G_{ij} + \eta_t^2 G_{ij}^2,$$

where $h_t^2$ and $\eta_t^2$ denote the proportions of phenotypic variance explained by additive genetic effects, and additive-by-additive interactions, respectively. In Eqn. (2.1), $c_t^2$ is the residual correlation that is not explained by IBD sharing between siblings (e.g., caused by shared environmental effects), and $G_{ij}$ is the realised relationship coefficient between siblings. As in **Supplementary Note 1**, we assume that $G_{ij} \sim N(0.5, s^2)$, where $s \approx 0.038$.

We now assume that these nonlinear effects are ignored in the analysis such that the model used to estimate heritability assumes that

(2.2) $$\mathrm{E}(y_i y_j | G_{ij}) = c^2 + h^2 G_{ij}$$

Under Eqn. (2.2), the expectation of the ordinary least-squares (OLS) estimators of $h^2$ (denoted $h_{\mathrm{OLS}}^2$) can be expressed as

(2.3) $$h_{\mathrm{OLS}}^2 = \frac{\mathrm{cov}(G_{ij}, y_i y_j)}{\mathrm{var}[G_{ij}]} = \frac{\mathrm{E}[\mathrm{E}(y_i y_j | G_{ij}) G_{ij}] - E[G_{ij}]E[y_i y_j]}{\mathrm{var}[G_{ij}]} = \frac{\left(c_t^2 E[G_{ij}] + h_t^2 E[G_{ij}^2] + \eta_t^2 E[G_{ij}^3]\right) - r_p/2}{\mathrm{var}[G_{ij}]}$$

where $r_p = c_t^2 + h_t^2/2 + \eta_t^2(1/4 + s^2)$ is the phenotypic correlation between siblings. Assuming that $G_{ij}$ is normally distributed, we have that

(2.4) $$E[G_{ij}^3] = E[G_{ij}]^3 + 3E[G_{ij}]\mathrm{var}(G_{ij}) = 1/8 + (3/2)s^2$$

Therefore, the numerator in Eqn. (2.3) can be rewritten as

(2.5) $$\mathrm{cov}(G_{ij}, y_i y_j) = \left[\frac{c_t^2}{2} + h_t^2\left(\frac{1}{4} + s^2\right) + \eta_t^2\left(\frac{1}{8} + \frac{3s^2}{2}\right)\right] - \frac{1}{2}\left[c_t^2 + \frac{h_t^2}{2} + \eta_t^2\left(\frac{1}{4} + s^2\right)\right] = (h_t^2 + \eta_t^2)s^2$$

which implies that $h_{\mathrm{OLS}}^2 = h_t^2 + \eta_t^2$. In other words, additive and additive-by-additive interactions effects are confounded, as previously shown by Young and colleagues.[4]

It also follows that the expectation of OLS estimator of $c^2$ (denoted $c_{\mathrm{OLS}}^2$) can be expressed as

(2.6) $$c_{\mathrm{OLS}}^2 = r_p - \frac{1}{2}h_{\mathrm{OLS}}^2 = c_t^2 + \frac{h_t^2}{2} + \eta_t^2\left(\frac{1}{4} + s^2\right) - \frac{1}{2}(h_t^2 + \eta_t^2) = c_t^2 - (1 - 4s^2)\eta_t^2/4$$
$$\approx c_t^2 - \frac{\eta_t^2}{4} < c_t^2$$

This last equation shows that not accounting for additive-by-additive interactions can lead to under-estimate $c_t^2$. Consistently, unmodelled non-additive genetic effects can also lead to negative $c_{\mathrm{OLS}}^2$ estimates.

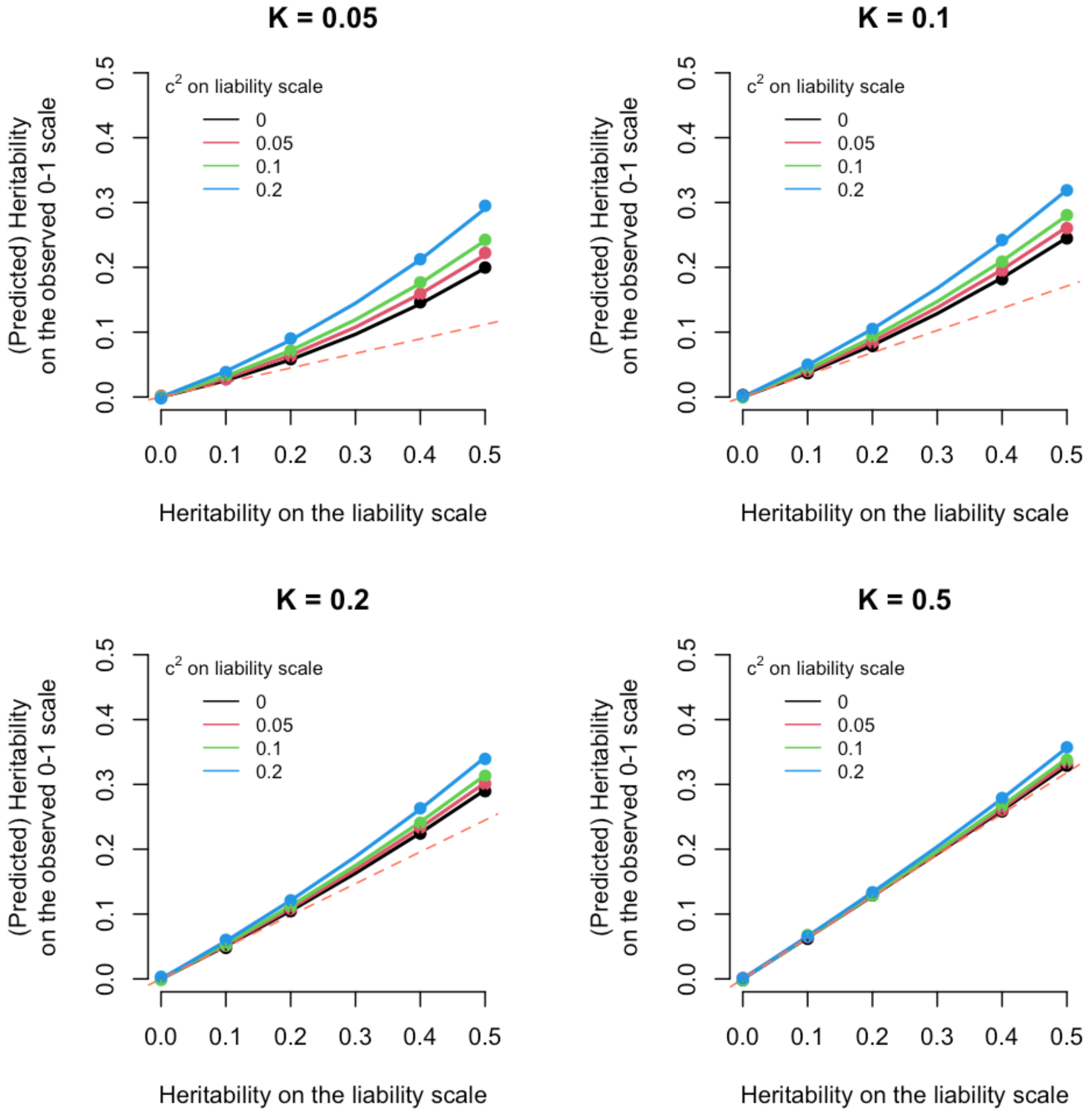


**Fig. S1. Theoretical relationship between $h_\ell^2$ (x-axis) and $h_{01}^2$ (y-axis) for different combinations of $(c_\ell^2, h_\ell^2)$ and $K$ as predicted by Eqn. (1.11).** The dotted line represents the line $y = \phi(t)^2/[K(1-K)]$, which corresponds to the linear transformation classically used for small relatedness values or small values of heritability. Each dot represents the average Haseman-Elston estimate over 30 replicates from 10,000,000 sibling pairs simulated under the corresponding parameter.

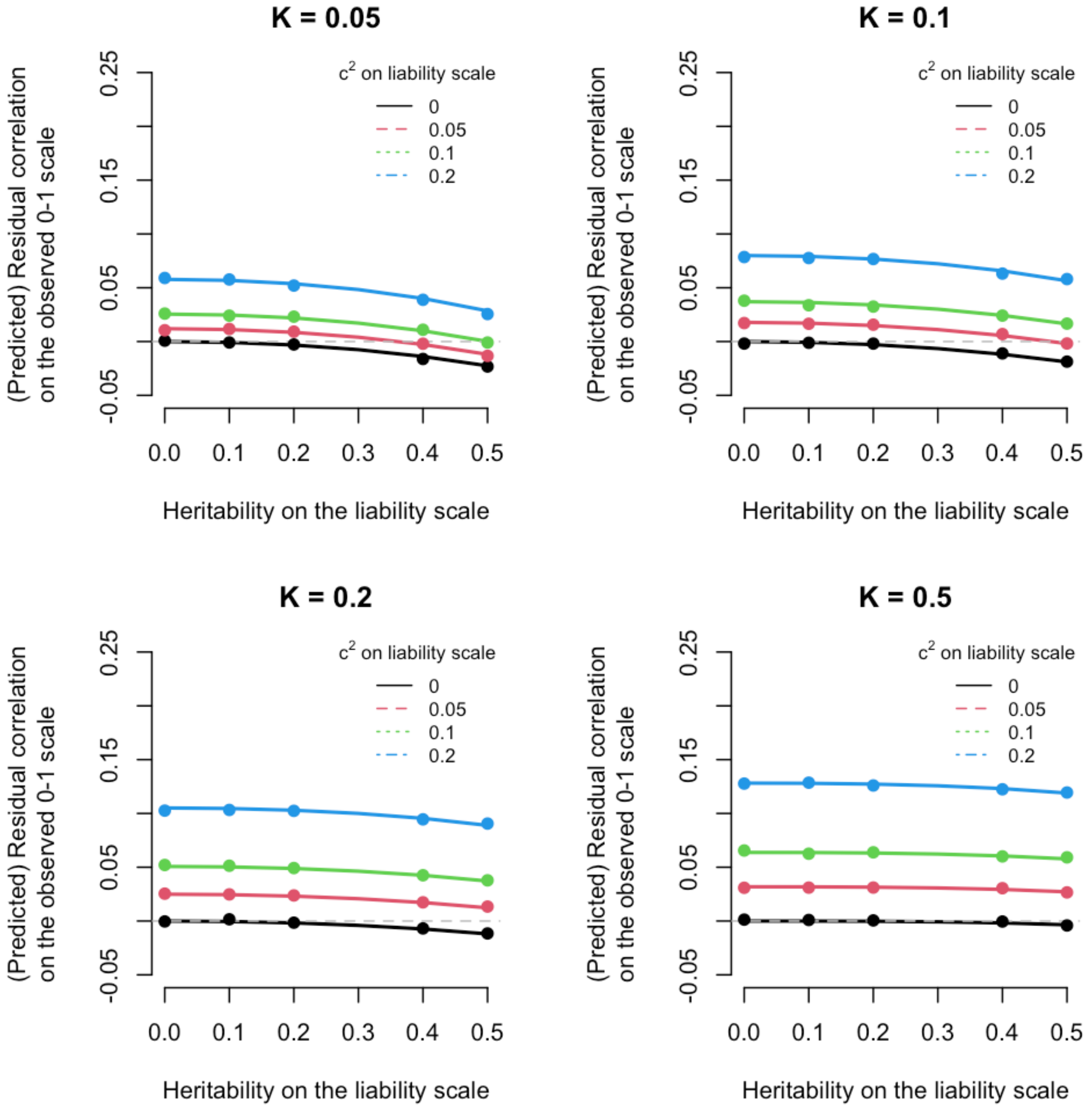


**Fig. S2. Theoretical relationship between $h_\ell^2$ (x-axis) and $c_{01}^2$ (y-axis) for different combinations of $(c_\ell^2, h_\ell^2)$ and $K$ as predicted by Eqn. (1.12).** Each dot represents the average Haseman-Elston estimate over 30 replicates from 10,000,000 sibling pairs simulated under the corresponding parameter.

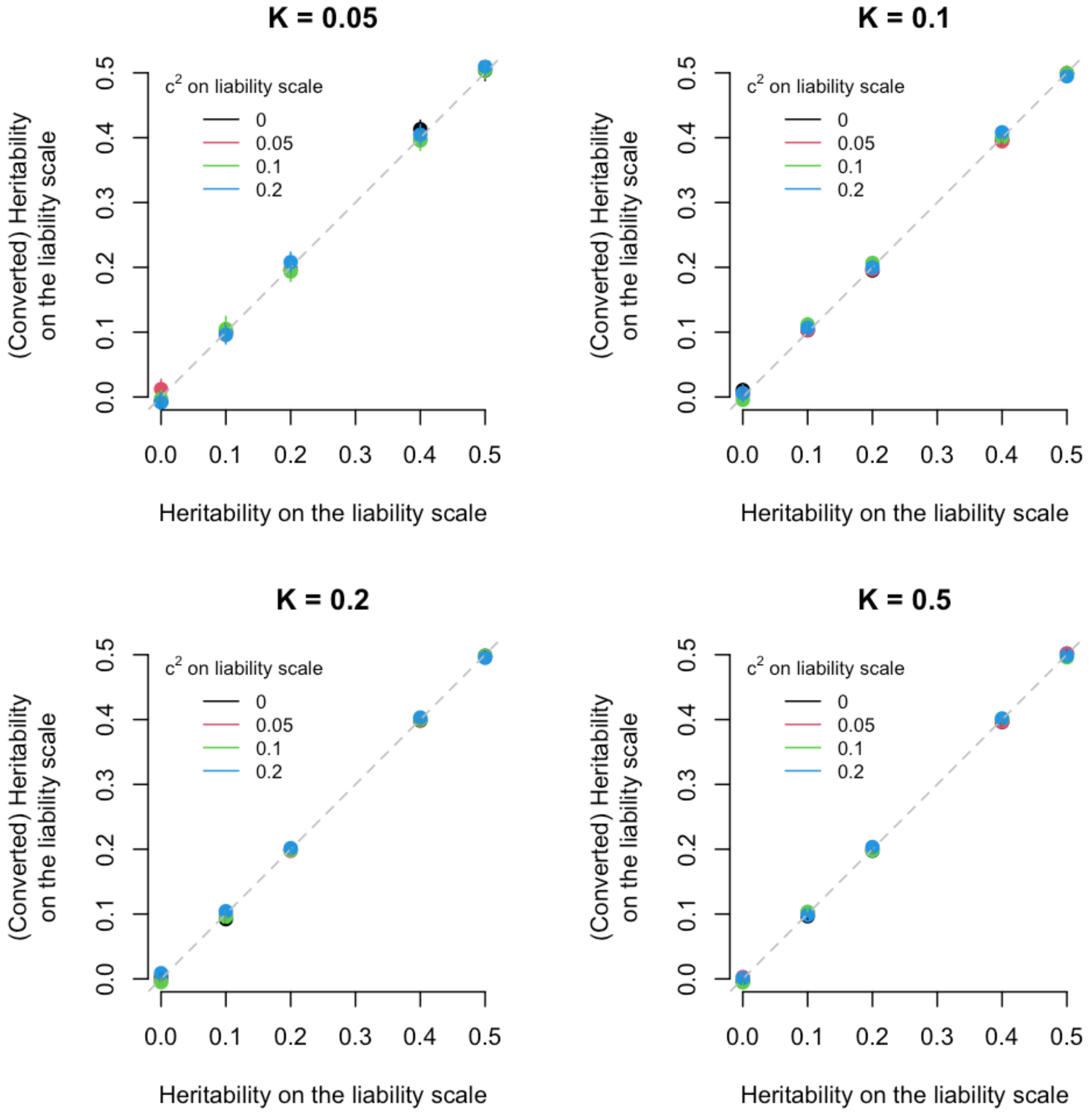


**Fig. S3. Relationship between expected (x-axis) and estimated (y-axis) $h_\ell^2$.** The parameter $h_\ell^2$ was estimated on the observed 0-1 scale then transformed back to the liability scale by solving Eqn. (1.13) numerically. Each dot represents the average Haseman-Elston estimate over 30 replicates from 10,000,000 sibling pairs simulated under the corresponding parameter.

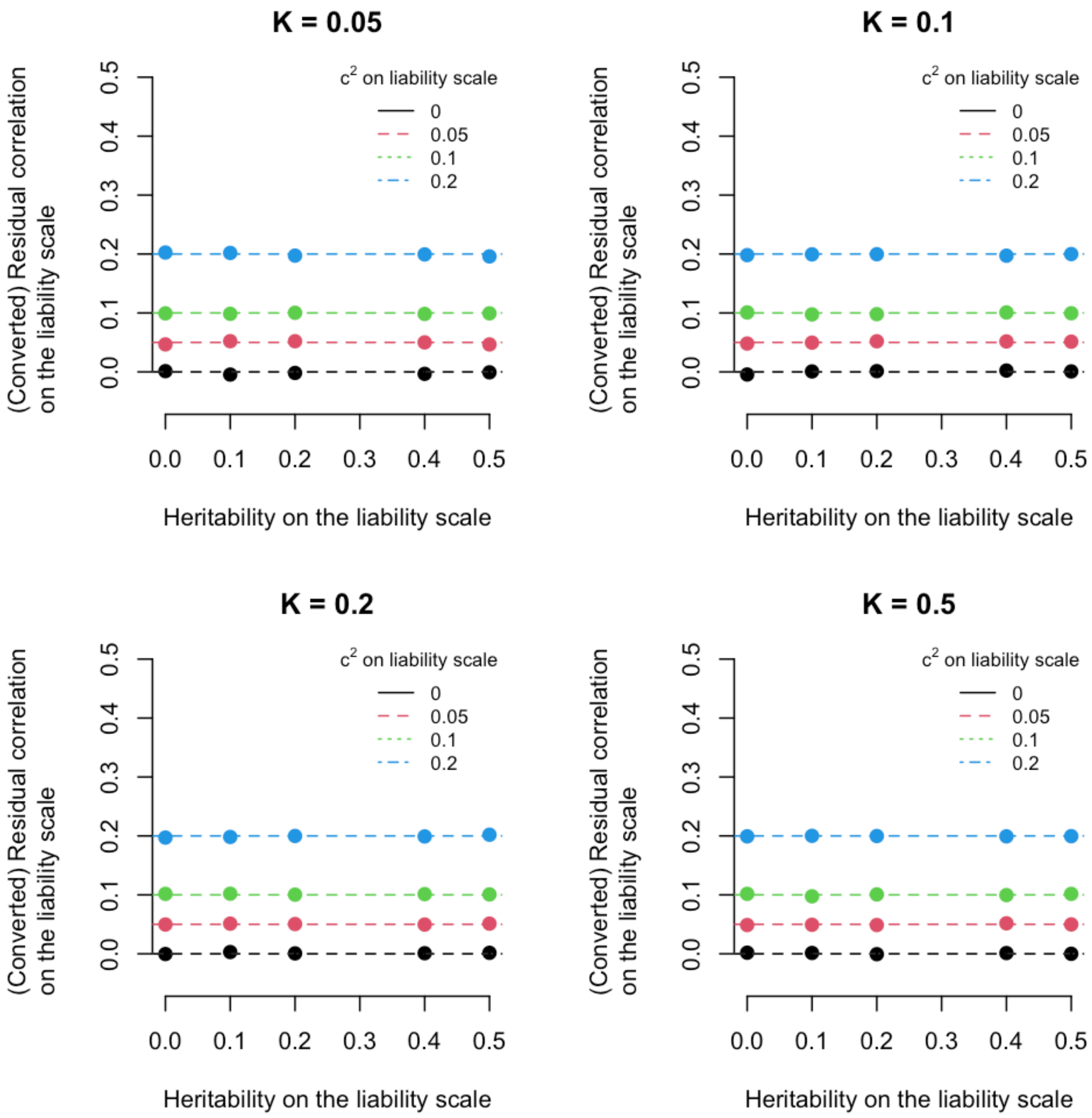


**Fig. S4. Relationship between expected (x-axis) and estimated (y-axis) $c_\ell^2$.** The parameter $c_\ell^2$ was estimated on the observed 0-1 scale then transformed back to the liability scale by solving Eqn. (1.13) numerically. Each dot represents the average Haseman-Elston estimate over 30 replicates from 10,000,000 sibling pairs simulated under the corresponding parameter.
